# Wastewater sample site selection to estimate geographically-resolved community prevalence of COVID-19: A research protocol

**DOI:** 10.1101/2020.08.23.20180224

**Authors:** Ray A. Yeager, Rochelle H. Holm, Kumar Saurabh, Joshua L Fuqua, Daymond Talley, Aruni Bhatnagar, Ted Smith

## Abstract

**Background:** Wastewater monitoring for virus infections within communities can complement conventional clinical surveillance. Currently, most SARS-CoV-2 testing is performed during clinical encounters with symptomatic individuals, and therefore likely underrepresents actual population prevalence. Randomized testing on a regular basis to estimate population-level infection rates is prohibitively costly and is hampered by a range of barriers associated with participation in clinical research. In comparison, community-level fecal monitoring can be performed through wastewater surveillance and can effectively surveil communities with less temporal lag than other surveillance methods. However, epidemiologically-defined protocols for wastewater sample site selection are lacking.

**Methods:** Herein we describe methods for developing a geographically-resolved population-level wastewater sampling approach in Jefferson County, Kentucky which may have general applicability for cities throughout the United States. This approach was developed by the selection of sampling locations along sewer lines transporting raw wastewater from geographically and demographically distinct areas that correspond with locations where random testing of residents occurs.

**Conclusions:** Development of this protocol for population-level sampling for SARS-CoV-2 prevalence in wastewater can be utilized to inform consistent wastewater monitoring among cities for up-to-date and geographically-resolved information on COVID-19 prevalence within communities. This information could substantially supplement public health surveillance of COVID-19 and thus serve to better guide targeted mitigation strategies throughout the United States.

## Background

Wastewater monitoring can complement clinical surveillance for global virus infections when fecal shedding has been established for the viral strain (Bisseux et al., 2020). For example, the application of wastewater epidemiology has been used for wide-range monitoring including illicit drug use, poliomyelitis, Hepatitis E, Human Immunodeficiency Virus, adenoviruses, and coronavirus for low-cost, real-time estimates, and retrospective monitoring (Bibby and Peccia 2013; Masclaux et al., 2013; Castiglioni et al., 2014; Pogka et al., 2017). Currently, COVID-19 clinical diagnostic testing is largely focused on symptomatic individuals; however, this underrepresents prevalence (Moghadas et al., 2020), and limiting detection to symptomatic individuals makes it difficult to reconstruct clusters of infection in a timely fashion through the aggregation of clinical and occupational diagnostic testing.

Fecal shedding of SARS-CoV-2 has been documented in patients diagnosed with COVID-19 (Wolfel et al, 2020). It is unsurprising, then, that SARS-CoV-2 is detectable in sewer systems, which could be an important early indicator of COVID-19 in communities (Ahmed et al., 2020a; Bivins et al., 2020; Medema et al., 2020; Peccia et al, 2020; Randazzo et al., 2020). However, interpretation of the signal, beyond presence/absence, of copies of SARS-CoV-2 RNA, independent of the direct sampling of known COVID-19 patients that contribute to that waste stream, continues to challenge broad application. Whereas a close relationship between stool and wastewater samples has been reported for poliomyelitis (Pogka et al., 2017), this is less well-established for SARS-CoV-2 (Kitajima et al., 2020; Wölfel et al., 2020). Because viral RNA monitoring of wastewater is done passively and anonymously, it may be an important option for community-level monitoring with the added benefit that it includes pre-symptomatic/asymptomatic individuals, underserved populations with less access to testing resources, and others who experience other barriers to clinical testing (Bivins et al., 2020). Moreover, wastewater sampling may also provide geographic location of infection clusters (“hot spots”) that are much more difficult to detect with conventional in-person measurements.

Wastewater monitoring of SARS-CoV-2 is available to complement community clinical diagnostic surveillance for infections at three units of analysis: 1) for an entire community at an established centralized wastewater facility; 2) for neighborhoods, within communities at sewer line manholes or pump stations; and 3) for congregate living facilities (e.g. ships, buildings) at effluent access points. Much of the published literature on the monitoring of SARS-CoV-2 in wastewater addresses samples collected at treatment facilities (Ahmed et al., 2020b; Medema et al., 2020). Bisseux et al. (2020) report the usefulness of combining raw wastewater sampling with clinical encounters in an urban area. From an epidemiological perspective, sampling derived from hundreds of thousands of households may produce useful aggregate infection prevalence information and may provide advance early warning about impending clinical facilities’ burden or the need for adjusting broad social restrictions. However, this approach does not address the possibility that important changes in infection dynamics in component areas of the community could trigger spatially-focused attention; examples could include public health intervention measures such as ad hoc “pop up” community testing or development of place- and culture-specific health communications and public service announcements.

To the best of our knowledge, there is no published methodological framework for sample site selection to maximize the surveillance and response value of SARS-CoV-2 wastewater testing within communities. Hence, we propose a pragmatic location-based method for selection of raw wastewater sample site locations, where wastewater testing results are representative of geographically-resolved community prevalence of SARS-CoV-2 infection. Such an approach could serve to inform public health officials and decision-makers on the comparative prevalence of SARS-CoV-2 infection within communities as well as inform proactive place-specific public health interventions.

Moving from central treatment facilities to neighborhood sewer lines introduces factors that preclude comparisons across sites. For example, sewer systems are not always accurately mapped, and sewer lines, in addition to household wastewater, may contain combined commercial, industrial, and often rain water, which complicates the consistent and comparable sampling of excreta due to inconsistent dilution of household waste. Furthermore, these lines often have complex technical and geographic characteristics from ad hoc expansion due to growing populations. Therefore, a key barrier to developing epidemiologically-comparable and geographically-resolved monitoring of SARS-CoV-2 in wastewater is the lack of established methods for sample site selection specifically designed for community-wide disease surveillance.

Based on sanitary sewer system design and function, a survey of sub-population viral load in wastewater involves raw samples taken directly from locations along sewer lines transporting water from geographically distinct areas. These raw wastewater sites are closer to individual sewer users and thus have a higher specificity than sludge and wastewater samples collected from treatment facilities, which degrade more over a longer travel time (GPEI, 2015). Shorter travel times have been recommended for SARS-CoV-2 samples (Hart and Halden, 2020).

Sampling of centralized treatment facilities is traditionally completed as part of environmental regulatory compliance monitoring of the effluent, prior to release to a waterway. At centralized treatment facilities, cycle times present an upper limit to how many individuals might possibly contribute to settled combined solids. For most urban treatment plants, solids will turn over every 24 hours. Gaining certainty regarding the temporal representativeness of the sample is a critical parameter. One conventional approach is to establish a pooled or composite sample, which is typically achieved by using an automated sampling device that takes regularly timed grabbed samples which are combined over 24 hours or some other interval. For example, the recommendation for wastewater monitoring of poliomyelitis is a single composite 24-h sample, taken weekly for one to three years (GPEI, 2015). The temporal dynamics of sampling frequency directly relate to the clinical actionability of the data obtained, as low frequency of aliquots will have low specificity to actual concentrations. For COVID-19, necessary frequency has yet to be empirically established for neighborhood scale surveillance. Given the likely variations in residential outflow, dilution, and mixing rate, frequent aliquots will be necessary for a representative 24-hour sample.

We developed the method we describe here in conjunction with a larger community COVID-19 prevalence assessment research project conducted in Jefferson County, KY (the Co-Immunity Project). The method supports SARS-CoV-2 surveillance in wastewater for community monitoring. Our main objective is to describe the establishment of geographically-resolved urban community surveillance for SARS-CoV-2 in wastewater. Our secondary aims are to: 1) identify wastewater field sampling techniques and limitations present in field conditions typical of urban settings which may require modified sampling site locations; and 2) describe scenarios for use of data from wastewater to inform public health interventions.

## Methods

As a component of a longitudinal COVID-19 community prevalence study in Jefferson County, KY, USA, (the Co-Immunity Project) at the University of Louisville, we additionally analyzed wastewater for prevalence of SARS-CoV-2. With a population of approximately 770,000, covering 380 square miles, Jefferson County is physically and demographically representative of many urban areas across the United States. There are five Water Quality Treatment Centers (WQTC) operated by Louisville/Jefferson County Metropolitan Sewer District (MSD), each treating wastewater from an average of 3.5 to 350 million gallons per day. We based our selection of sample media on the sanitary sewer system design and absence of WQTCs for sampling of sludge with consistent geographic resolution of areas with fewer than 100,000 residents. Though the WQTCs are fixed, we have continuously added and removed sampling sites to refine our sampling approach during the pilot. Based on this approach and field sampling limitations (Figure 1), we selected a total 15 sample sites: 10 community sites collecting from manholes and pump stations and five WQTCs (Figure 2).

**Figure 1:**
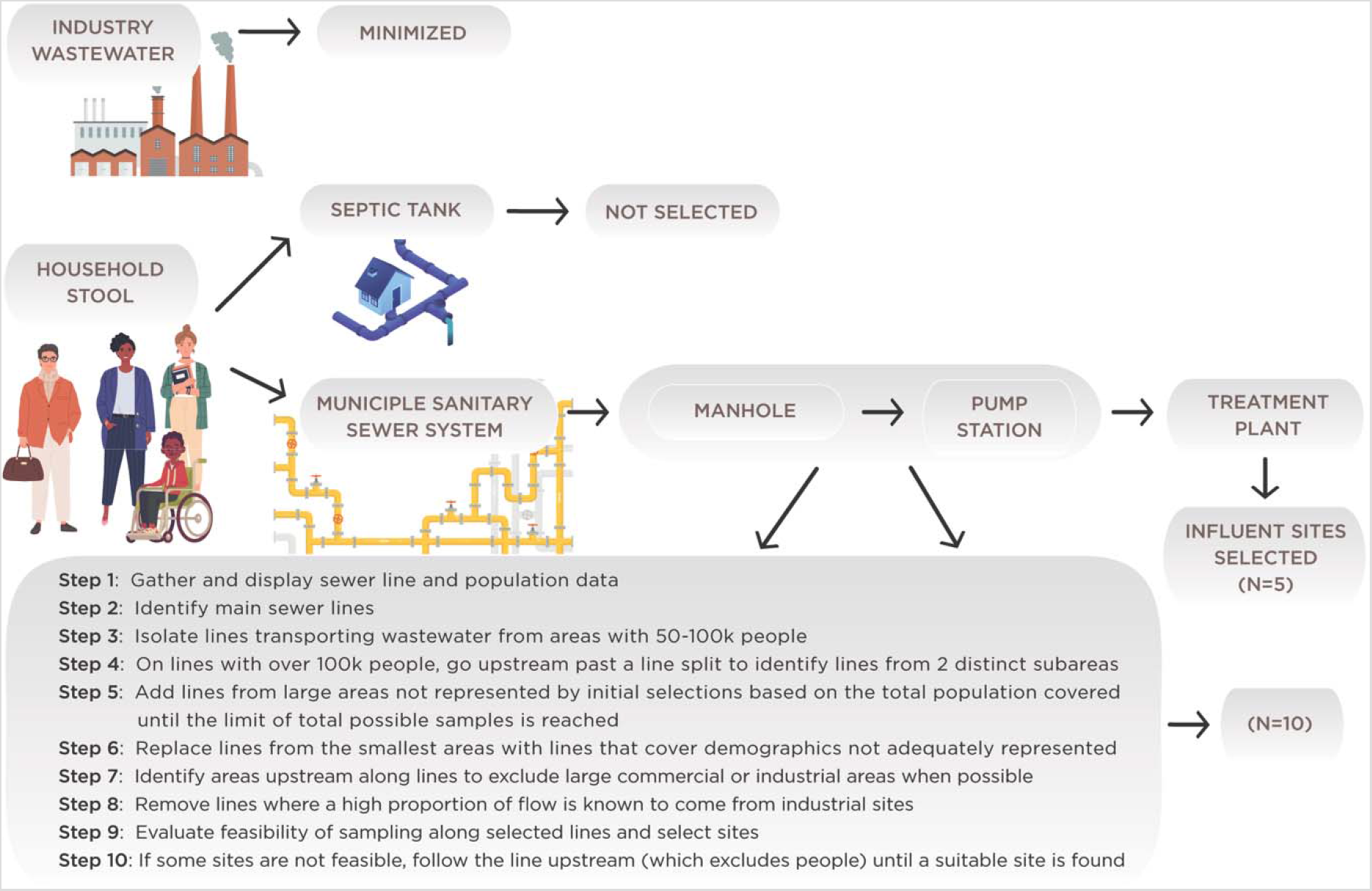
Flow chart representing the protocol for selection of monitoring sites for SARS-CoV-2 in wastewater

**Figure 2:**
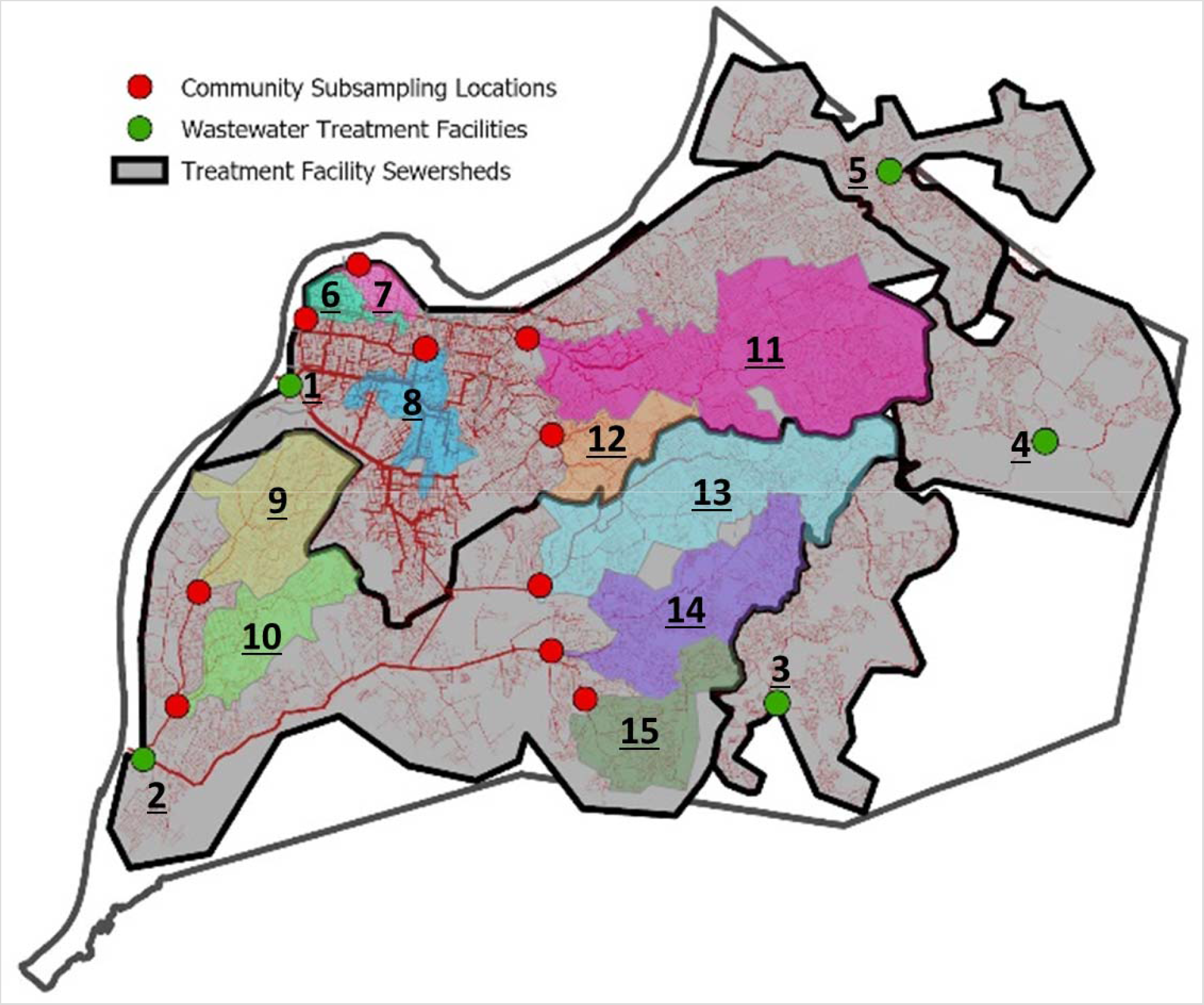
Location of wastewater sampling sites and corresponding catchment areas in Jefferson County, Kentucky

### Data

We collected existing sewer data in Geographic Information Systems (GIS) format from the MSD, allowing for rapid design and deployment of this protocol. These data included information on the location and attributes of all sewer lines and access points within the city. Citywide socioeconomic data was collected from the U.S. Census Bureau, including information on the distribution of income, race, ethnicity, and population distribution characteristics.

### Sample size justification

In Jefferson County, sludge samples would not allow a geographically-resolved understanding of SARS-CoV-2 infection rates and would be limited to monitoring the five WQTCs in the city. The recommendation for wastewater poliomyelitis monitoring is a sample population of 100,000 to 300,000 persons (GPEI, 2015); however, our project required sample sensitivity to be directly comparable with our concurrent nasal diagnostic clinical study, justifying a smaller sample population at urban section or neighborhood resolution related to zip code populations with raw wastewater. Based on the specific design of sewer lines in Jefferson County, there was a steep decline in informative value of additional sites beyond these 15 locations.

### Ethics

For wastewater surveillance, we over-selected geographic areas that are typically underrepresented in clinical testing. These are often low-income and communities of color. The research team has discussed this new approach to SARS-CoV-2 surveillance in community meetings and in local media interviews, and it typically is met with supportive interest. Wastewater samples cannot be used to identify individuals and are considered “community samples” by the sewer district; thus the sampling is not subject to institutional review for human studies.

Rationale for choosing sampling locations and field sampling limitations

- Inclusion criteria:

- Residential coverage with water toilets serviced by the municipal sanitary sewer system
- Locations overlapping recruitment areas for randomized resident testing of the corresponding prevalence study
- Anonymized SARS-CoV-2 concentration data in wastewater with a minimum population of 5,000
- Accessibility to sewer lines with free-flowing wastewater (street lines, pumping stations, and main treatment facilities)
- Contextual information (flow rate, temperature)
- Feasibility with field human resources
- For manhole locations, free-flowing wastewater at no more than 25 feet below the street level due to sampler equipment limitations
- Exclusion criteria

- Downstream of a majority commercial and industrial sewer outflow industrial or hospital sites, even if surrounded by residential households
- Water toilets not connected to the municipal sanitary sewer system, such as areas where septic tanks are in use
- Any sites not practical for field sampling, such as the area around a manhole not offering enough space or safety for field personnel or traffic safety issues
- High temperature wastewater
- Specific additions

- Each WQTC
- Higher sampling in northwestern portion of the city to ensure inclusion of residents with disproportionate health risk

### Site selection

We identified ideal sample sites using a stepwise approach (Figure 1). Predominately residential areas were prioritized over large commercial areas due to predictable outflows from residential locations and the ability to precisely quantify area population and demographics with census and property data. We based demographic estimates on 2018 American Community Survey (ACS) block group data and aggregated to the sewersheds with overlapping block group centroids. In some cases, a large main sewer line was not accessible in areas with smaller feeder lines. Consequently, we moved sample sites upstream or sampled before two larger sewer lines joined. This was justified since, if sites were moved downstream, they would have overlapped zones that could not be geographically isolated. We then sent sample site recommendations to MSD, which reviewed these recommendations for site-specific practicality. Some practical obstacles included manholes that were not easily accessible for the sampling team (requiring 4-wheel drive vehicles) and manholes that were located along high traffic roadways. Contribution of hospitals or other facilities with high disinfectant effluent were minimized via our sampling approach.

We sampled each treatment facility in the county (green points in Figure 2), giving nearly 100% community coverage. The two major treatment facilities (Site ID 1 and 2) are downstream from some of the neighborhood sampling zones but also include neighborhood lines which were not included in community sampling catchment areas. This circumstance provides future opportunity to validate virus detection from feeder lines that are sampled and also infer signals from lines that were not sampled in some neighborhoods.

After our initial site selection, we performed an initial round of sampling. Upon field observations, we excluded one site due to higher than expected industrial outflow visually observed within the sample. Additionally, this sample was at an elevated temperature due to high temperature industrial discharges into the sewer system. Elevated temperatures have been reported to make wastewater samples vulnerable to over or under estimation due to possible degradation over time (Hart and Halden, 2020; Kitajima et al., 2020).

Specific accommodations for sample sites were also made based on field sampling logistics determined by MSD’s occupational safety standards for field activities. The rationale for choosing the number of sampling sites was also constrained by MSD staff availability, which in this case was two field teams who each required a full day to collect six to eight sample sites. Additionally, the work needed to be completed in time to prepare the samples for delivery and processing in the University of Louisville laboratory.

**Table 1.** Demographics of study sampling site catchment areas in Jefferson County, Kentucky^a^

| ID | Household income (USD\$) | | Race and Hispanic Origin | | |
| --- | --- | --- | --- | --- | --- |
|  | (mean within catchment |  | Non- |  |  |
|  | areas of reported block |  | Hispanic | Black | Hispanic |
|  | group median values) | Population | White (%) | (%) | (%) |
| 1 | 53,891 | 641,410 | 70 | 23 | 6 |
| 2 | 53,577 | 295,910 | 72 | 21 | 7 |
| 3 | 76,606 | 55,928 | 82 | 12 | 4 |
| 4 | 113,699 | 32,460 | 87 | 8 | 3 |
| 5 | 106,769 | 31,269 | 75 | 14 | 4 |
| 6 | 27,695 | 10,739 | 9 | 88 | 1 |
| 7 | 27,446 | 7,820 | 68 | 26 | 3 |
| 8 | 25,365 | 25,836 | 51 | 43 | 4 |
| 9 | 45,895 | 35,956 | 59 | 37 | 4 |
| 10 | 51,656 | 25,073 | 83 | 12 | 5 |
| 11 | 77,842 | 99,061 | 87 | 7 | 4 |
| 12 | 68,259 | 139,251 | 79 | 13 | 5 |
| 13 | 53,542 | 73,666 | 63 | 28 | 9 |
| 14 | 61,837 | 46,659 | 75 | 18 | 8 |
| 15 | 63,642 | 22,437 | 80 | 13 | 5 |

<sup>a</sup> Based on 2018 U.S Census Bureau American Community Survey (ACS)

Wastewater Sample Collection, Transport, Preservation, and Storage

Monitoring data include 24-h composite sample wastewater samples for SARS-CoV-2 analysis collected at least once per week. All samples are considered raw wastewater. We contracted with MSD for access and sample extraction at sample sites. Sampling personnel from MSD setup a portable automated composite autosampler (60–2954–001 Model GLS Sampler) the day prior to sampling. The sampler collects 30 ml into a 4L plastic container every 15 min. Ice was placed in the space between the cover and the collection jar as part of setting up the sampler. Transit time between different sites were less than 1 h to minimize differences that may lead to artifactual differences in virus count measurements, and the 15 samples were mostly collected within 5 h. This sampling approach minimizes variation in estimates of virus prevalence due to temperature fluctuations, even though viral degradation may vary because of temperature. Furthermore, population demographics will not change since we, consistently over time, collect samples from areas of known demographic characteristics.

In a large-scale field sampling program, an exact 24-h composite sample is difficult, but Schwartzbrod et al. (1985) suggest a 15-h minimum composite sample for evaluation of viral populations in wastewater. The time should be next day, as close to a full 24-h as possible. The next day, the sampling container was accessed. In some cases, it was located below a manhole and suspended with rope or on the ground adjacent to a sampling site. The sample was stirred by hand and a portable pump was subsequently used to fill two sample containers (one 500 ml and one 150 ml). Sampling personnel from MSD wore standard personal protective equipment(PPE) for wastewater sampling, including Tyvek coveralls, boots, hard hats, face shields, and gloves. After sampling, the portable pump and tubing were rinsed with bleach water and double rinsed with deionized water.

Although composite samples are preferred, evidence during our piloting found that grab samples would be required in instances of battery malfunction of the composite sampler or the composite sampler tubing being clogged by solid waste. To collect a grab sample, a sampling cup on a rope was used. Though grab sample data is still important, it was flagged for analysis and use in any final decision-making, due its incomplete representation of SARS-CoV-2 prevalence. Additionally, we used both 10L and 4L composite sample collection containers during the pilot The 4L containers were preferable to allow more volume of ice in the space between the cover and the collection jar. Additionally, our preferred set-up had the composite sampler placed securely at street/ground level, but in several neighborhood sample locations, for safety and security reasons, the composite sampler was lowered into the manhole and suspended. In that circumstance, the 4L sample collection container is preferable to reduce the weight of the sample that is manually raised to ground level.

Samples were transported on ice packs to the University of Louisville laboratory. One sample was stored at –60°C and one sample at 4°C for at least 5 h. The samples were split, with half retained for subsequent analysis at University of Louisville and the other half delivered to our academic lab for analysis. Samples were archived at least at –60°C for later analyses as needed (Dolfing, 2020). The further processing of these samples follows RNA extraction and detection using methods that are nearly identical to the methods applied to nasal swab samples. For a review of these see Ahmed et al. (2020c).

## Discussion

Here we propose a method to select locations for wastewater sampling for the purpose of geographically-resolved estimation of community prevalence of SARS-CoV-2 infection rates. The development of an epidemiologically-sound methodology for COVID-19 surveillance depends upon clear definition of the spatiotemporal properties of monitoring methods and clarity regarding the assumptions about the correspondence between the amount of RNA detected in different sampling configurations and the estimation of infected persons in that service area. In Jefferson County, monitoring the five WQTCs in the city would allow for nearly complete population surveillance, but it would not allow for an understanding of SARS-CoV-2 infection rates with high spatial resolution. Hence, we selected additional community-based wastewater sampling sites to allow for additional geographic resolution among WQTC catchment areas, utilizing a pragmatic approach to maximize public health value of our monitoring resources. To date, we are not aware of any other work using a similar methodology at the urban scale. While wastewater cannot be a substitute for clinical testing data, it may be utilized to estimate community-level infection rates, and may lead to new interventions within communities to contain emerging clusters of infection. This can also be done passively at a lower cost and with fewer human resources, making it a more feasible approach than regular widespread individual testing. The findings of this research strengthen the knowledge base to utilize wastewater monitoring in conjunction with clinical surveillance to provide critical insight to isolating and quantifying community-level SARS-CoV-2 infection prevalence and developing actionable public health responses.

This study contributes to advancement of knowledge by informing selection of sample sites for wastewater to correlate with spatial clinical data on infection rates from randomized testing. An important strength of this sampling approach is the provision of more accurate estimates of actual community-level SARS-CoV-2 infections, independent of the proportion of symptomatic individuals, individual testing capacity, healthcare capacity, socioeconomic capacity, and other substantial biases that skew prevalence estimates based on individual-level testing. Furthermore, sampling of wastewater closer to residences, as described here, allows for less time for virus to degrade before sampling, as opposed to centralized facility testing, thus allowing for more specific prevalence estimates than WQTC samples alone. The results of this geographically-resolved community testing approach provide a useful and cost-effective scenario for public health surveillance to inform interventions and countermeasures to allow geographically-targeted strategies to reduce transmission of SARS-CoV-2.

There are several limitations to this protocol. The site selection is framed to consider physical variation in sample state (travel time, sample temperature, flow data, background chemistry, variation in shedding by infected persons); however, the rapidly evolving understanding of decay of SARS-CoV-2 in wastewater may further refine sample location selection. In some cases, sewer line access was restricted. The number of sites was not predetermined, but based on spatial resolution of sewer catchment areas and on physical limitations of the system to select ideal population distributions. Flow data was not included in the same site selection criteria. In some areas, MSD has a combined sewer and storm water system, possibly diluting residential sources of SARS-CoV-2 or introducing non-toilet sources of SARS-CoV-2. However, dilution may largely be accounted for by normalizing to population and flow rate, as well as biological measurements of fecal dilution markers. Further, while we can partially account for resulting dilution, it is not possible to fully exclude industrial and commercial outflow. It should be noted that clinical data from testing the community’s residents potentially includes households not represented in the sanitary sewer system (e.g., septic tank users, individuals living in one area but spending the majority of their day in another area of the city).

Areas for future exploration include: 1) sensitivity and temporal dynamics to determine if raw wastewater is a concurrent indicator of SARS-CoV-2 measurement in sludge; 2) sensitivity and community limits of detection including frequency of sampling or moving sites to determine community trends; 3) quantification of field samples such as turbidity or a rapid field proxy for SARS-CoV-2 in sewers; 4) waste travel time in relation to sampling location selections; 5) attributes of physical sewer system infrastructure regarding age of sewage or opportunities for latent virus in the system; and 6) the need for sewer sampling infrastructure that is “pandemic ready” and a better fit design for answering human health surveillance instead of only industrial surveillance.

## Conclusion

Development of this protocol for population-level sampling for SARS-CoV-2 prevalence in wastewater can be utilized to inform consistent wastewater monitoring among cities for up-to-date and geographically-resolved information on COVID-19 prevalence within communities. This information could substantially supplement public health surveillance of COVID-19 and thus serve to better guide targeted mitigation strategies throughout the United States.

## Data Availability

The authors confirm that the data supporting the findings of this study are available within the article [and/or] its supplementary materials.

## Declarations

### Ethics approval and consent to participate

Not applicable.

### Consent for publication

Not applicable.

### Availability of data and materials

The data are available upon request. For expression of interest, please contact (Ted Smith).

### Authors’ contributions

Conceptualization: R.A.Y, D.T., A.B. and T.S.; Methodology R.A.Y, D.T., A.B. and T.S; Writing-original draft preparation: R.H.H.; Writing-review and editing: R.A.Y, R.H.H., K.S., J.L.F, D.T., A.B. and T.S.; Supervision: A.B. and T.S.; Project administration: T.S. All authors have read and agreed to the published version of the manuscript.

## Acknowledgements

We thank our field and analytical technicians Dwight Mitchell, David Hoetker, Rick Strehl, and Ian Santisteban for their diligent commitment to this work.

